# Multistrain probiotics ameliorate tau pathology and preserve visuospatial cognition in early cognitive impairment: A double-blind, randomized controlled trial

**DOI:** 10.64898/2026.02.01.26345102

**Authors:** Eun Hyun Seo, Sarang Kang, Seung-Gon Kim, Ja-Hae Kim, Hye-Jung Yoon, Kyu Yeong Choi, Hyung-Jun Yoon, Kun Ho Lee, Kang-Ho Choi

## Abstract

**Background:** Emerging evidence suggests that microbiota play a role in Alzheimer’s disease (AD) pathology and cognitive performance. Interventions targeting the oral–brain axis may offer neuroprotective benefits, particularly during the early stages of cognitive impairment. This randomized controlled trial (RCT) investigated whether a multistrain probiotic supplement could modulate AD-related plasma biomarkers and cognitive function in older adults with early cognitive impairment.

**Methods:** Participants from the Gwangju Alzheimer’s Disease and Related Dementia (GARD) Cohort in Korea were enrolled in a double-blind, randomized, placebo-controlled trial. Older adults with early cognitive impairment were randomized to receive either a multistrain probiotic supplement (KL-P301) or a placebo for 24 weeks. Plasma pTau181, glial fibrillary acidic protein (GFAP), and neurofilament light chain (NfL) were quantified at baseline and follow-up. Cognitive and clinical assessments included the Clinical Dementia Rating (CDR), Mini-Mental State Examination (MMSE), CERAD neuropsychological battery, Stroop test, and Rey–Osterrieth Complex Figure (ROCF). Treatment effects were analyzed using paired t-tests, linear mixed models, and ANCOVA adjusted for baseline and demographic covariates.

**Results:** Of the 87 participants analyzed (probiotic, n=40; placebo, n=47), the probiotic group exhibited a significant reduction in plasma pTau181 levels compared with the placebo group (*p* < 0.001). While GFAP and NfL levels remained stable in the probiotic group, the placebo group showed significant longitudinal increases (*p* = 0.014 and *p* = 0.041, respectively). Clinically, the probiotic group demonstrated improved CDR (*p* = 0.010), primarily driven by the memory domain. Domain-specific cognitive analyses revealed that the probiotic group significantly improved in visuospatial construction (Constructional Praxis, *p* = 0.036; ROCF copy, *p* = 0.027) and maintained stable constructional recall, whereas the placebo group showed a significant decline (*p* = 0.025). No significant between-group differences were observed in MMSE, verbal memory, or executive/attentional functions.

**Conclusion:** The multistrain probiotic supplement reduced tau-related pathology and neuroinflammation-associated biomarkers and selectively preserved visuospatial construction and visual memory in older adults with early cognitive impairment. These findings suggest that modulating the oral-immune–brain axis with multistrain probiotics represents a viable, non-pharmacological strategy to slow AD-related pathological progression and cognitive decline in early-stage patients.

## Background

Preventing Alzheimer’s disease (AD) at a very early stage, before extensive neurodegeneration can occur, has become a primary goal of dementia research [1]. Early intervention during the preclinical phase has the greatest potential to delay disease progression. Thus, developing new early preventive strategies focusing on modifiable biological and lifestyle factors has become a central goal in AD research [1, 2]. The gut–immune–brain axis is a bidirectional signaling network linking the gut microbiota with the brain via neural, hormonal, and immune pathways [3, 4]. Recently, O’Riordan et al. [3] indicated that the immune system serves as a key mediator in this axis, regulating neuroinflammatory responses, microglial activity, and cognitive function through microbial signaling. This integrative perspective implies that probiotics may act beyond simple anti-inflammatory effects, helping restore the balance of the disrupted microbiota–immune–brain network and potentially influencing amyloid-β, tau pathology, and cognitive function.

In particular, accumulating evidence indicates that alterations in the gut microbiota contribute to AD pathology via multiple interconnected mechanisms. Gut dysbiosis has been shown to increase gut permeability and activate immune responses, which may weaken the blood–brain barrier and contribute to neuroinflammation and neuronal damage in AD [5]. Moreover, probiotic interventions can attenuate amyloid-β accumulation and tau pathology, as well as improve cognitive performance in transgenic AD mouse models [6–8]. Collectively, these findings indicate that maintaining or restoring the gut microbial balance through probiotic supplementation may represent a promising strategy to mitigate early amyloid and tau pathology and preserve cognitive function during the prodromal stage of AD [3].

Among the pathological hallmarks of AD, abnormal tau phosphorylation and aggregation show the closest association with early cognitive decline and disease progression [9]. Notably, associations of both visuospatial ability and visual episodic memory with tau accumulation have been reported in numerous studies [10–13]. These visuospatial cognitive functions are particularly sensitive to tau pathology and therefore represent valuable markers for tracking changes in disease status. Given the growing evidence that the gut–brain axis may modulate neuroinflammatory and tau-related processes, probiotic interventions could potentially mitigate the downstream effects of tau accumulation and preserve visuospatial function in the prodromal stage of AD. However, despite these mechanistic links, it remains unclear whether multistrain probiotic supplementation specifically targeting the oral microbiota can modulate AD-related tau pathology and domain-specific cognitive function in older adults with early cognitive impairment.

Among various microbial taxa, lactic acid bacteria (LAB) and *Bifidobacterium* species have been widely recognized for their anti-inflammatory, antioxidant, and neuroprotective properties in both animal and human studies [4]. Especially, a mixture of probiotic bacteria Bifidobacterium animalis ssp. lactis, Limosilactobacillus fermentum, and Levilactobacillus brevis have shown promising biological activity in previous studies. For example, Lee et al. [14] demonstrated that this probiotic combination alleviated LPS-induced cognitive impairment in mice through anti-inflammatory and antioxidant mechanisms, significantly reducing circulating IL-1β and nitric oxide levels while upregulating antioxidant enzymes. Subsequently, these findings were further extended to a transgenic Alzheimer’s model, showing that the same probiotic mixture could attenuate amyloid-β accumulation, tau phosphorylation, and microglial activation in 5xFAD mice [6].

Therefore, based on these cumulative results, the present study employed the same three-strain combination of *Bifidobacterium animalis ssp. lactis, Limosilactobacillus fermentum,* and *Levilactobacillus brevis* as a biologically validated probiotic formulation with anti-inflammatory, antioxidant, and neuroprotective potential relevant to cognitive and neurodegenerative disorders. We aimed to investigate the effects of this specific multistrain probiotic supplement targeting the oral microbiota on AD-related pathological biomarkers and cognitive function in older adults with early cognitive impairment, using a 24-week, double-blind, randomized controlled trial.

## Methods

### Study Design

The study was a double-blind, randomized, placebo-controlled, single-center clinical trial conducted at the Department of Neurology, Biomedical Research Institute of Chonnam National University Hospital (Seoul, Republic of Korea), from April to December 2023. The study included a 24-week period with five scheduled visits: screening (Visit 1), baseline (Visit 2), and three follow-up visits during the intervention period (Visit 3 at Week 4, Visit 4 at Week 12, and Visit 5 at Week 24 or early termination). Written informed consent was obtained from all participants at the screening visit. Demographic information, medical and medication histories, and concomitant drug use were also assessed. Vital signs, anthropometric measurements, and laboratory tests were further conducted to confirm eligibility. The participants were also screened for adverse reactions and evaluated for suitability. At baseline (Visit 2), eligible participants underwent APOE genotyping, comprehensive clinical and neuropsychological assessment, self-report questionnaires, and blood biomarker tests. During the intervention phase, the participants received either multistrain probiotics or a placebo daily for 24 weeks. Follow-up visits were conducted at Week 4 and Week 12 to monitor compliance, record adverse events, measure vital signs, and repeat laboratory safety assessments. Cognitive function and biomarker evaluations were repeated at the final visit (Week 24) to assess efficacy and safety. Safety assessments included clinical laboratory tests, urinalysis, physical examination, and monitoring of treatment-emergent adverse events (TEAEs). Compliance was verified at each visit by counting returned supplementary packages and confirming adherence.

### Interventions

The probiotic intervention used in the present study was a multistrain probiotic supplement targeting the oral microbiota. The active intervention (code name: KL-P301) consisted of a probiotic formulation containing three strains: *Bifidobacterium animalis ssp. lactis KL101, Limosilactobacillus fermentum KL271,* and *Levilactobacillus brevis KL251*. Participants consumed one sachet orally once daily, either directly or combined with water. The placebo (KL-P301 placebo) was identical in appearance and packaging to the active product, but contained maltodextrin rather than probiotic strains. Participants consumed one sachet orally once daily in the same manner as the active product. Both products were manufactured by Kolab Inc. (Gwangju, Republic of Korea) and stored at room temperature, with a shelf life of 12 months from the manufacturing date.

### Randomization and blinding

Eligible participants were randomly assigned to either the active or placebo group in a 1:1 ratio. A randomization table was independently generated and maintained by the Contract Research Organization (CRO), MEDITIP (Total Medical Service Consulting, http://meditip.co.kr). All investigators, participants, and study personnel were blinded to allocation throughout the trial. The randomization code remained sealed and opened only after the database was locked. The active and placebo products were identical in appearance, color, and packaging to maintain blinding. Throughout the study, investigators, coordinators, clinical research associates, and data managers remained unaware of group assignments. Blinding was maintained until study completion. Code opening (unblinding) was performed by the CRO after database locking. The unblinding process and timing were both documented.

### Participants

We included individuals with early cognitive impairment recruited from the Gwangju Alzheimer’s Disease and Related Dementias (GARD) Cohort. The GARD database has been described in detail previously [15, 16]. Participants were included in the study if they: (1) were 60 years of age or older at screening; (2) were able to take oral medications; (3) had no severe hearing, visual, or language impairment that could interfere with neuropsychological testing; (4) had no dementia, history of stroke, alcohol or substance abuse, or other central nervous system disorders; (5) provided written informed consent prior to enrollment; (6) had evidence of cognitive complaints or decline, defined as a Mini-Mental State Examination (MMSE) score range 20 to 27; and (7) were judged to have subjective cognitive decline (SCD) or mild cognitive impairment (MCI) based on clinical interview by a neurologist. Participants were excluded if they: (1) had cognitive impairment due to vascular cognitive impairment or medical or psychiatric conditions (including epilepsy, acute neurological illness, major depressive disorder, bipolar disorder, schizophrenia, or were taking any related medications); (2) were using antibiotics at screening or expected to require them during the trial; (3) had a history of food poisoning within 4 months prior to screening; (4) were using or expected to use immunosuppressive agents during the study; (5) were considered to at risk of immunodeficiency; (6) were taking long-term psychiatric medications for depression or alcohol dependence; (7) had a history of head trauma with loss of consciousness; (8) had undergone cancer treatment within the past 3 years; (9) had implanted metallic devices; (10) were unable or unwilling to discontinue other probiotic products during the study; (11) were unable or unwilling to discontinue brain supplements or cognitive therapeutic agents during the study; (12) had a known allergy to the multistrain probiotics; (13) had participated in another interventional clinical trial within 3 months prior to screening or planned to participate in one during the study; and (14) were deemed otherwise unsuitable for participation by a neurologist. This study was approved by the Institutional Review Board of Chonnam National University Hospital (IRB no: CNUH-2023-072). Written informed consent to participate was obtained from each participant or their legal guardian. The study was conducted in accordance with the principles of the Declaration of Helsinki.

### Clinical and neuropsychological assessments

All participants were interviewed and accordingly assigned a Clinical Dementia Rating (CDR) by a neurologist with advanced training in dementia research. The Geriatric Depression Scale (GDS)[17], a self-report measure, was administered. Comprehensive neuropsychological assessments were administered by experienced psychologists. These assessment included the MMSE [18], Rey-Osterrieth Complex Figure (ROCF) test, comprising the Copy, Immediate recall, and Delayed recall; the Stroop Color and Word Test (SCWT) comprising the Word page, Color page, and Color-Word page subtests [19]; and the Consortium to Establish a Registry for Alzheimer’s Disease neuropsychological battery (CERAD-NP), comprising the Verbal fluency (VF), 15-item Boston Naming Test (BNT), Word List Memory (WLM), Word List Recall (WLR), Word List Recognition (WLRc), Constructional Praxis (CP), Constructional Recall (CR), and Trail Making Test (TMT) A and B subtests[20].

### Blood biomarker and amyloid burden positivity

Blood samples were collected from study participants. All collected blood samples were subsequently processed and separated according to a standardized protocol to obtain cell-free plasma. The plasma biomarkers, specifically phosphorylated Tau at threonine 181 (pTau181), phosphorylated Tau 217 (pTau217), Glial fibrillary acidic protein (GFAP), and neurofilament light chain (NfL), were quantified using the highly sensitive Single Molecule Array (Simoa) platform. The following commercial kits from Quanterix were utilized for quantification: the pTau-181 Advantage V2.1 Kit (104111) for pTau181, the ALZpath pTau 217 Advantage PLUS assay kit (104570) for pTau217, and the Neurology 2-plex kit (103520) for GFAP and NfL. All assays were performed according to the manufacturer’s instructions.

Amyloid burden positivity was determined using 18F-florbetaben positron emission tomography (PET) at baseline. All participants underwent imaging according to standardized acquisition protocols. Visual assessment was performed across key cortical regions (frontal, parietal, temporal, and posterior cingulate), and each scan was rated using the Brain β-Amyloid Plaque Load (BAPL) system [21]. Based on this scoring scheme, scans with a BAPL score of 1 were classified as amyloid-negative, whereas scores of 2 or 3 were categorized as amyloid-positive.

### Data Control Process

All clinical and neuropsychological data were collected using paper case report forms (CRFs), rather than electronic CRFs (eCRFs). Investigators recorded the results of the cognitive assessments and questionnaires on paper CRFs, and clinical research associates (CRAs) verified their accuracy against the source documents. Following completion of the study, the CRO’s data management team digitized all CRF data and performed range and logic checks to identify any errors or inconsistencies. Queries were issued to the investigators for resolution, and all corrections were documented. Once all queries were resolved and data integrity was confirmed, the database was locked, with approval from both the sponsor and the principal investigator. After database locking, no further data modification was permitted, and the finalized datasets (ITT, PP, Safety sets) were analyzed according to a predefined statistical analysis plan.

### Statistical analysis

All statistical analyses were conducted using SPSS software (version 29.0.1.0; IBM Corp., Armonk, NY, USA) with complete-case data, without imputation. Baseline demographic and clinical characteristics were compared between the control and probiotic groups using independent t-tests for continuous variables and chi-square tests for categorical variables. Within-group changes from baseline to week 24 were assessed using the paired t-test. Between-group differences in the change scores were examined using independent t-tests. To compare group differences at week 24 while adjusting for potential confounders, an analysis of covariance (ANCOVA) was conducted, with age, sex, education level, and baseline value for each outcome included as covariates. Furthermore, a group × baseline interaction term was included to evaluate whether any baseline levels moderated between-group differences at follow-up. For outcomes reported as percentiles or z-scores, only baseline values were adjusted without any further adjustment for age, sex, or education. For variables requiring post-hoc pairwise comparisons, Fisher’s Least Significant Difference (LSD) test was applied. All statistical tests were two-tailed, and a p-value < 0.05 was considered statistically significant.

## Results

### Participant characteristics

Initially, 100 participants who met the inclusion and exclusion criteria were enrolled and randomized in the study. However, 13 participants (10 in the probiotic and 3 in the placebo groups) dropped out during the intervention period (Figure 1). The reasons for dropout were as follows: contraindicated medication use (four in the probiotic group and one in the placebo group), elective surgical treatment (two in the probiotic group), withdrawal of consent (two in the probiotic group and one in the placebo group), and mild adverse events (two in the probiotic group and one in the placebo group), which included nausea, edema, and diarrhea. The final analysis, therefore, used data from 87 participants (40 in the probiotic group and 47 in the placebo group). As shown in Table 1, the two groups were comparable with respect to all baseline demographic and clinical characteristics. The mean age of the total sample was 73.89 ± 5.2 years, with similar age distributions across groups (probiotic: 74 ± 4.8; placebo: 74 ± 5.1; *p* = 0.670). Gender distribution and education level showed no significant difference between the groups. The GDS scores were low in both groups, indicating the absence of any clinically meaningful depressive symptoms with no significant between-group differences. Cognitive status was also comparable, with mean MMSE scores of 25.35 ± 1.93 and 24.91 ± 1.83 in the probiotic and placebo groups, respectively (*p* = 0.284), with respective global CDR scores of 0.43 ± 0.18 and 0.46 ± 0.14 (*p* = 0.350). The proportion of APOE ε4 carriers was similar between groups (47.5% vs. 57.4%; *p* = 0.354), and amyloid positivity also did not differ significantly between the placebo and probiotic groups (63.8% vs. 50.0%, respectively; *p* = 0.193), indicating that baseline genetic risk and amyloid burden were comparable across groups. Baseline plasma biomarkers, including pTau217, GFAP, and NfL, were also similar between the groups. Overall, no statistically significant baseline differences were observed between the groups, indicating that randomization was balanced.

**Figure 1.**
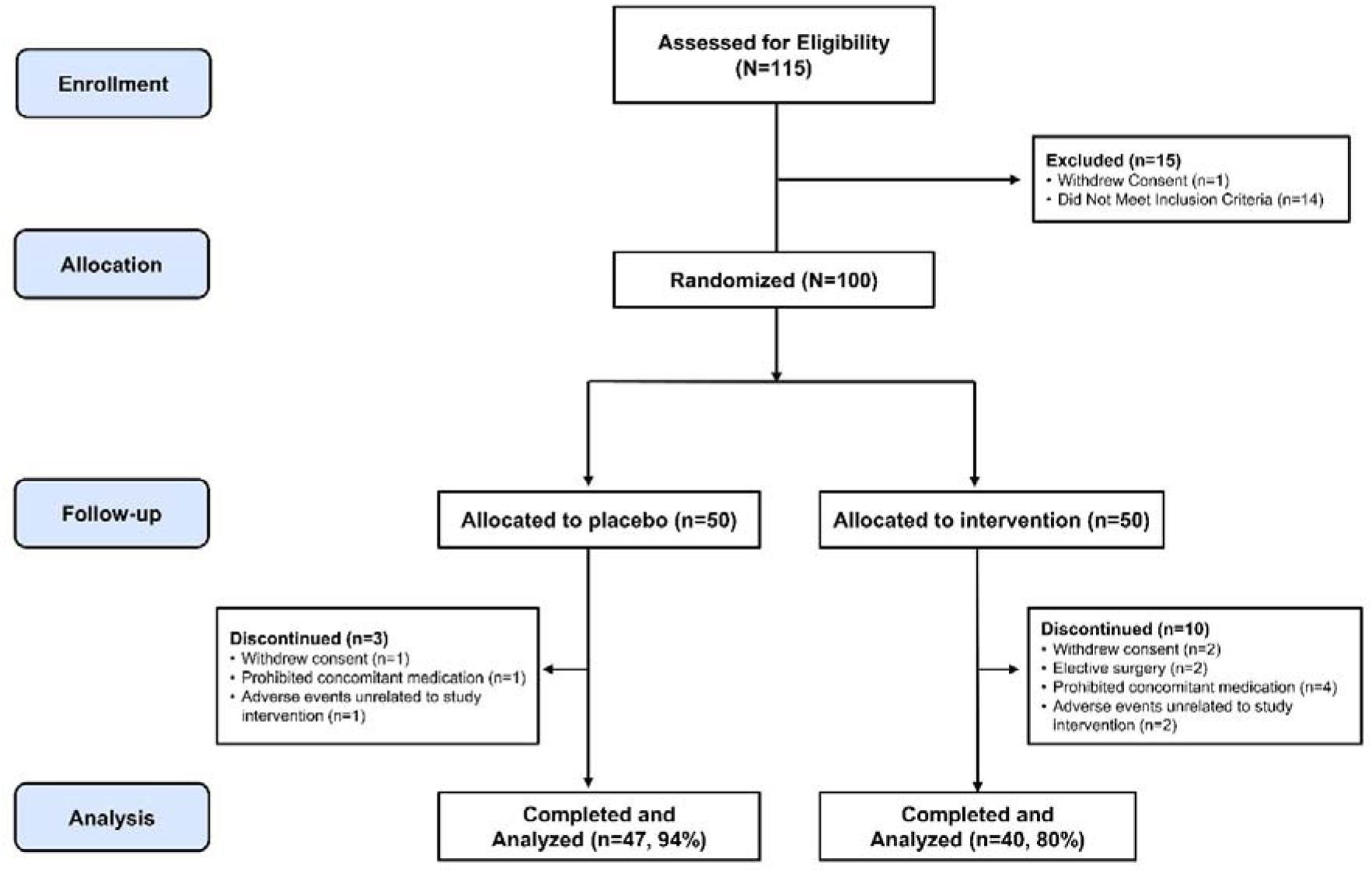
Summary of patient flow. Flow diagram depicting participant progression through the randomized, double-blind clinical trial.

**Table 1.** Baseline demographic and clinical characteristics of the study participants.

|  | Placebo group | Probiotic group | Total | *P*-value |
| --- | --- | --- | --- | --- |
|  | (*n* = 47) | (*n* = 40) | (*n* = 87) |  |
| Age, y | 74 (5.1) | 74 (4.8) | 73.89 (5.2) | 0.670 |
| Sex, *n* (%) |  |  |  |  |
| Female | 33 (70.21) | 28 (70) | 61 (70.11) | 0.583 |
| Male | 14 (29.79) | 12 (30) | 26 (29.89) |  |
| Education, y | 9.02 (3.66) | 8.5 (4) | 8.77 (3.85) | 0.620 |
| MMSE | 24.91 (1.83) | 25.35 (1.93) | 25.11 (1.88) | 0.284 |
| GDS, | 8.55 (6.11) | 9.50 (7.47) | 8.73 (6.43) | 0.517 |
| CDR | 0.46 (0.14) | 0.43 (0.18) | 0.44 (0.16) | 0.350 |
| APOE carrier, *n* (%) | 27 (57.4) | 19 (47.5) | 46 (52.9) | 0.354 |
| Amyloid+, *n* (%) | 30 (63.8) | 20 (50.0) | 50 (57.5) | 0.193 |
| pTau181, pg/mL | 22.20 (7.83) | 24.52 (10.08) | 23.26 (8.58) | 0.845 |
| pTau217, pg/mL | 0.58 (0.40) | 0.65 (0.42) | 0.61 (0.40) | 0.784 |
| GFAP, pg/mL | 212.13 (92.03) | 196.32 (78.84) | 204.86 (74.65) | 0.220 |
| NfL, pg/mL | 14.95 (4.74) | 15.56 (5.83) | 18.18 (29.1) | 0.490 |

Values are presented as the mean (SD) or number (%)
Abbreviations: MMSE, mini-mental state examination; GDS, Geriatric Depression Scale; Amyloid+, amyloid burden positive determined by positron emission tomography; CDR, clinical dementia rating; APOE, Apolipoprotein E; pTau181, phosphorylated tau 181; GFAP, glial fibrillary acidic protein; NfL, neurofilament light chain

### Clinical and neuropsychological function

Across the 24-week intervention period, the probiotic group demonstrated a significant improvement in the CDR Sum of Boxes (SB) compared to the placebo group (*p* < 0.05; Figure 2A). This effect was notably driven by the memory domain, with no significant differences observed in other domains (Table 2). In contrast, the placebo group showed no significant changes in either the CDR global score or any domain-specific scores. Neither group demonstrated significant changes in GDS scores over time.

**Figure 2.**
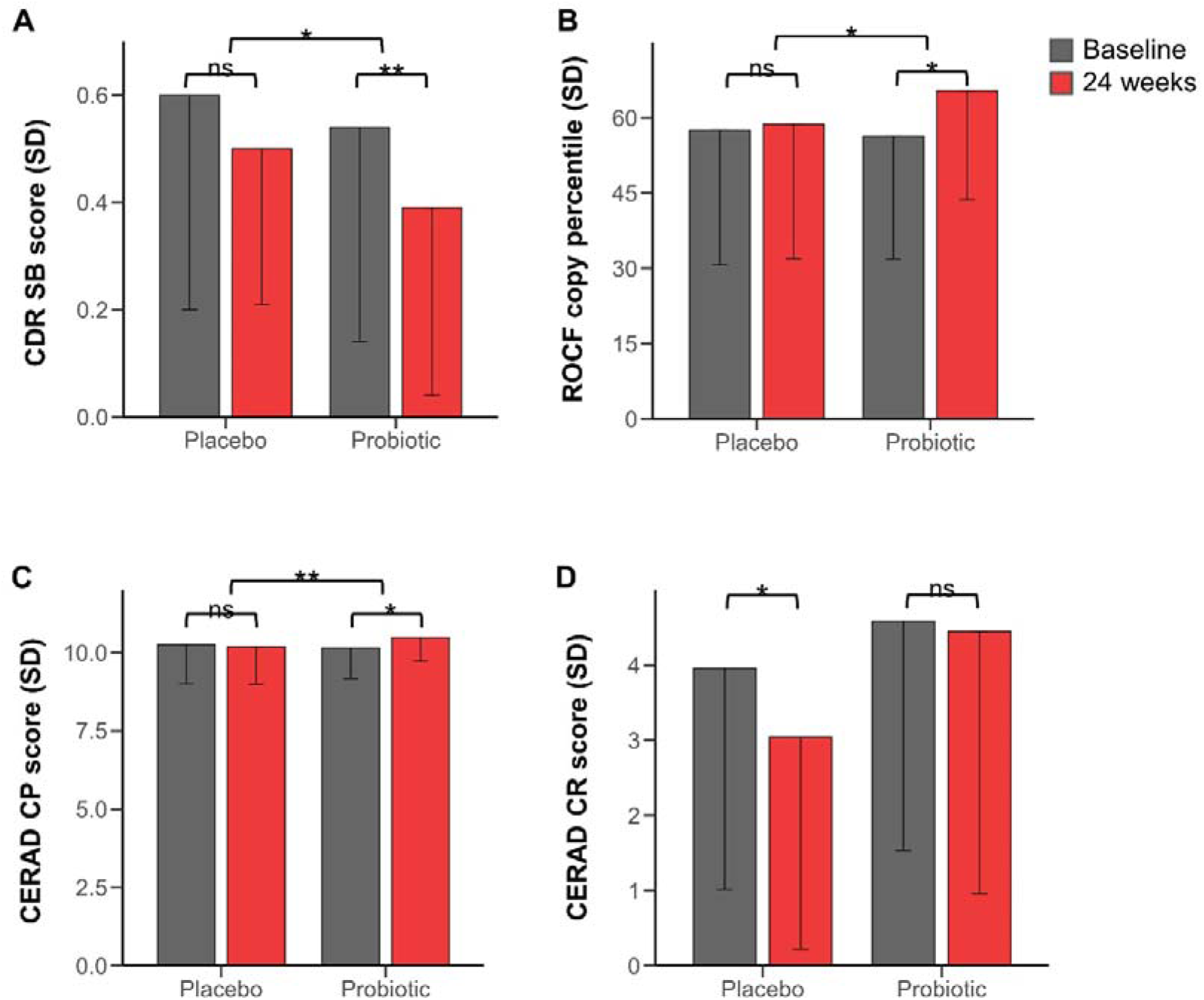
Changes in cognitive test from baseline to 24-weeks in the probiotic and placebo groups. Bar plots (A-F) presenting pre-intervention (baseline, shown in black) and post-intervention (24-week, shown in red) scores for CDR. Group differences and within group changes are annotated using significance (*p<0.05, **p<0.01, ns=not significant). Abbreviations: CDR, clinical dementia rating; ROCF copy: Rey-Osterrieth complex figure test copy; CERAD: Consortium to Establish a Registry for Alzheimer’s Disease; CP: Constructional Praxis; CR: Constructional Recall

**Table 2.**
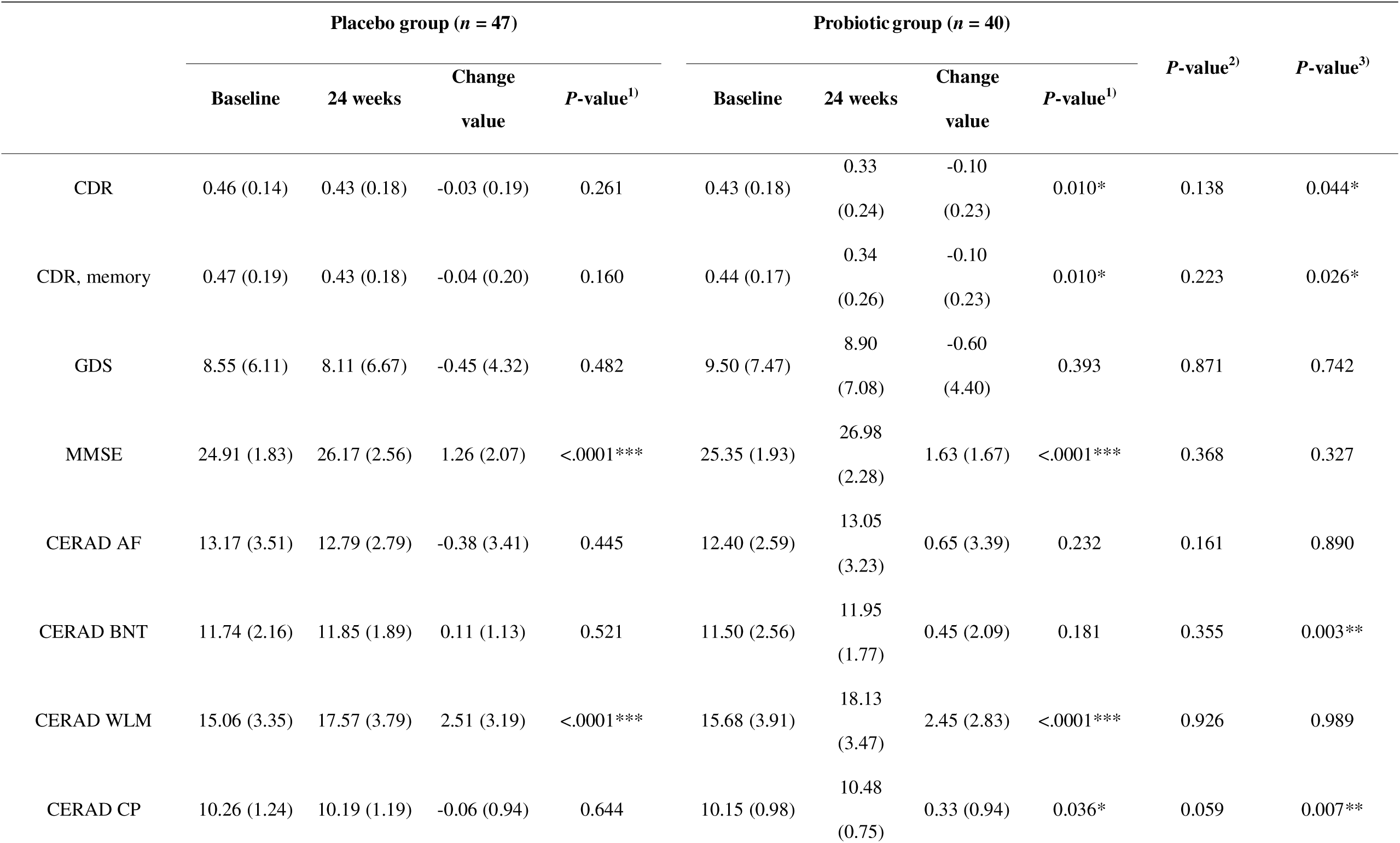
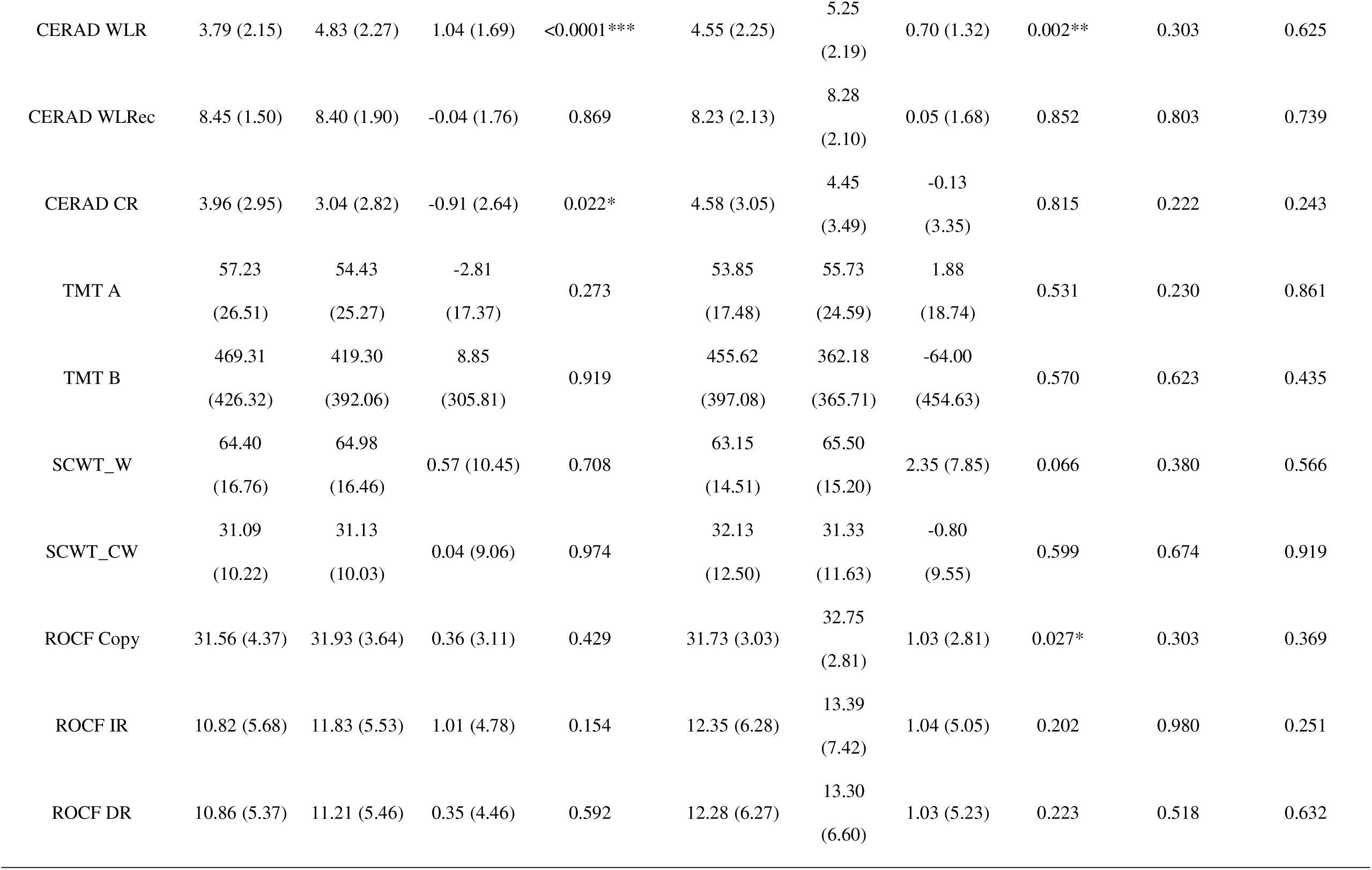
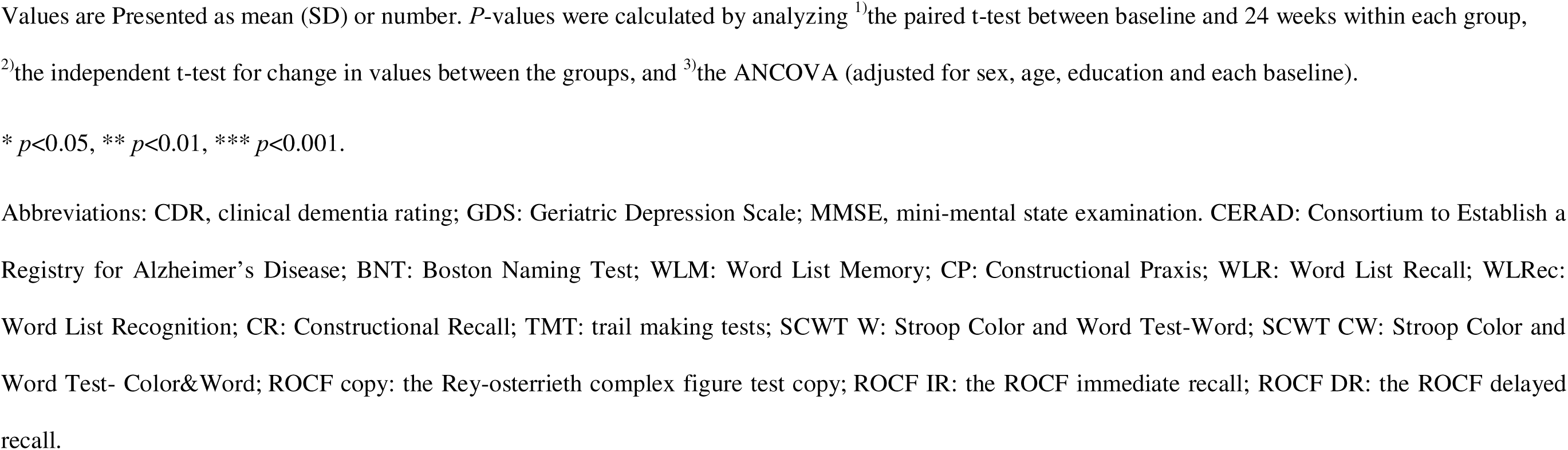
Group difference in clinical and neuropsychological measures.

|  | Placebo group (*n* = 47) |  |  |  | Probiotic group (*n* = 40) |  |  |  | *P*-value<sup>2)</sup> | *P*-value<sup>3)</sup> |
| --- | --- | --- | --- | --- | --- | --- | --- | --- | --- | --- |
|  | Baseline | 24 weeks | Change value | *P*-value<sup>1)</sup> | Baseline | 24 weeks | Change value | *P*-value<sup>1)</sup> |  |  |
| CDR | 0.46 (0.14) | 0.43 (0.18) | -0.03 (0.19) | 0.261 | 0.43 (0.18) | 0.33 (0.24) | -0.10 (0.23) | 0.010* | 0.138 | 0.044* |
| CDR, memory | 0.47 (0.19) | 0.43 (0.18) | -0.04 (0.20) | 0.160 | 0.44 (0.17) | 0.34 (0.26) | -0.10 (0.23) | 0.010* | 0.223 | 0.026* |
| GDS | 8.55 (6.11) | 8.11 (6.67) | -0.45 (4.32) | 0.482 | 9.50 (7.47) | 8.90 (7.08) | -0.60 (4.40) | 0.393 | 0.871 | 0.742 |
| MMSE | 24.91 (1.83) | 26.17 (2.56) | 1.26 (2.07) | <.0001*** | 25.35 (1.93) | 26.98 (2.28) | 1.63 (1.67) | <.0001*** | 0.368 | 0.327 |
| CERAD AF | 13.17 (3.51) | 12.79 (2.79) | -0.38 (3.41) | 0.445 | 12.40 (2.59) | 13.05 (3.23) | 0.65 (3.39) | 0.232 | 0.161 | 0.890 |
| CERAD BNT | 11.74 (2.16) | 11.85 (1.89) | 0.11 (1.13) | 0.521 | 11.50 (2.56) | 11.95 (1.77) | 0.45 (2.09) | 0.181 | 0.355 | 0.003** |
| CERAD WLM | 15.06 (3.35) | 17.57 (3.79) | 2.51 (3.19) | <.0001*** | 15.68 (3.91) | 18.13 (3.47) | 2.45 (2.83) | <.0001*** | 0.926 | 0.989 |
| CERAD CP | 10.26 (1.24) | 10.19 (1.19) | -0.06 (0.94) | 0.644 | 10.15 (0.98) | 10.48 (0.75) | 0.33 (0.94) | 0.036* | 0.059 | 0.007** |
| CERAD WLR | 3.79 (2.15) | 4.83 (2.27) | 1.04 (1.69) | <0.0001*** | 4.55 (2.25) | 5.25<br>(2.19) | 0.70 (1.32) | 0.002** | 0.303 | 0.625 |
| CERAD WLRec | 8.45 (1.50) | 8.40 (1.90) | -0.04 (1.76) | 0.869 | 8.23 (2.13) | 8.28<br>(2.10) | 0.05 (1.68) | 0.852 | 0.803 | 0.739 |
| CERAD CR | 3.96 (2.95) | 3.04 (2.82) | -0.91 (2.64) | 0.022* | 4.58 (3.05) | 4.45<br>(3.49) | -0.13<br>(3.35) | 0.815 | 0.222 | 0.243 |
| TMT A | 57.23<br>(26.51) | 54.43<br>(25.27) | -2.81<br>(17.37) | 0.273 | 53.85<br>(17.48) | 55.73<br>(24.59) | 1.88<br>(18.74) | 0.531 | 0.230 | 0.861 |
| TMT B | 469.31<br>(426.32) | 419.30<br>(392.06) | 8.85<br>(305.81) | 0.919 | 455.62<br>(397.08) | 362.18<br>(365.71) | -64.00<br>(454.63) | 0.570 | 0.623 | 0.435 |
| SCWT_W | 64.40<br>(16.76) | 64.98<br>(16.46) | 0.57 (10.45) | 0.708 | 63.15<br>(14.51) | 65.50<br>(15.20) | 2.35 (7.85) | 0.066 | 0.380 | 0.566 |
| SCWT_CW | 31.09<br>(10.22) | 31.13<br>(10.03) | 0.04 (9.06) | 0.974 | 32.13<br>(12.50) | 31.33<br>(11.63) | -0.80<br>(9.55) | 0.599 | 0.674 | 0.919 |
| ROCF Copy | 31.56 (4.37) | 31.93 (3.64) | 0.36 (3.11) | 0.429 | 31.73 (3.03) | 32.75<br>(2.81) | 1.03 (2.81) | 0.027* | 0.303 | 0.369 |
| ROCF IR | 10.82 (5.68) | 11.83 (5.53) | 1.01 (4.78) | 0.154 | 12.35 (6.28) | 13.39<br>(7.42) | 1.04 (5.05) | 0.202 | 0.980 | 0.251 |
| ROCF DR | 10.86 (5.37) | 11.21 (5.46) | 0.35 (4.46) | 0.592 | 12.28 (6.27) | 13.30<br>(6.60) | 1.03 (5.23) | 0.223 | 0.518 | 0.632 |

Values are Presented as mean (SD) or number. *P*-values were calculated by analyzing <sup>1)</sup>the paired t-test between baseline and 24 weeks within each group, <sup>2)</sup>the independent t-test for change in values between the groups, and <sup>3)</sup>the ANCOVA (adjusted for sex, age, education and each baseline).
\* $p < 0.05$ , \*\* $p < 0.01$ , \*\*\* $p < 0.001$ .
Abbreviations: CDR, clinical dementia rating; GDS: Geriatric Depression Scale; MMSE, mini-mental state examination. CERAD: Consortium to Establish a Registry for Alzheimer's Disease; BNT: Boston Naming Test; WLM: Word List Memory; CP: Constructional Praxis; WLR: Word List Recall; WLRec: Word List Recognition; CR: Constructional Recall; TMT: trail making tests; SCWT W: Stroop Color and Word Test-Word; SCWT CW: Stroop Color and Word Test- Color&Word; ROCF copy: the Rey-osterrieth complex figure test copy; ROCF IR: the ROCF immediate recall; ROCF DR: the ROCF delayed recall.

Regarding neuropsychological measures, the two groups showed distinct patterns of change across the selected subtests (Table 2). Performances on two tests assessing visuospatial construction ability were significantly improved in the probiotic group (*p* = 0.027 for ROCF copy; *p* = 0.036 for CERAD CP; Figure 2B and 2C). However, the placebo group showed no significant changes in either test. Furthermore, ANCOVA revealed significant group differences in CERAD CP score (p=0.007) and ROCF copy (z-score, *p* = 0.037). The CERAD CR score in the probiotic group remained stable (*p* = 0.815), whereas that in the placebo group declined significantly (*p* = 0.025) over the 24-week intervention period (Figure 2D). Further mixed model analysis of the CR score revealed a significant group–time interaction (*p* < 0.0001).

However, both groups showed significant improvement on several tests, including the MMSE and CERAD verbal memory scores, with no significant difference in the magnitude of change between the groups. No significant changes were observed in either group for the remaining cognitive assessments, including AF, BNT, TMT A & B, Stroop test scores, and ROCF memory scores, and no significant inter-group differences were found for these measures.

### AD and neurodegeneration biomarker

As shown in Table 3, administration of the multistrain probiotic supplement demonstrated significant positive effects on plasma biomarkers of AD after 24 weeks compared to the control group. Plasma pTau181 levels significantly decreased in the probiotic group at 24 weeks, whereas no significant change was observed in the placebo group (Figure 3A). This group difference was the most statistically robust (*p* < 0.001). Plasma pTau217 levels did not change significantly in either group during the 24-week intervention (Figure 3B). GFAP and NfL levels remained stable relative to baseline in the probiotic group during the 24-week intervention period. Conversely, the placebo group exhibited a significant increase in both GFAP (*p* = 0.014) and NfL levels (*p* = 0.041). over the same period (Figure 3 C and D).

**Figure 3.**
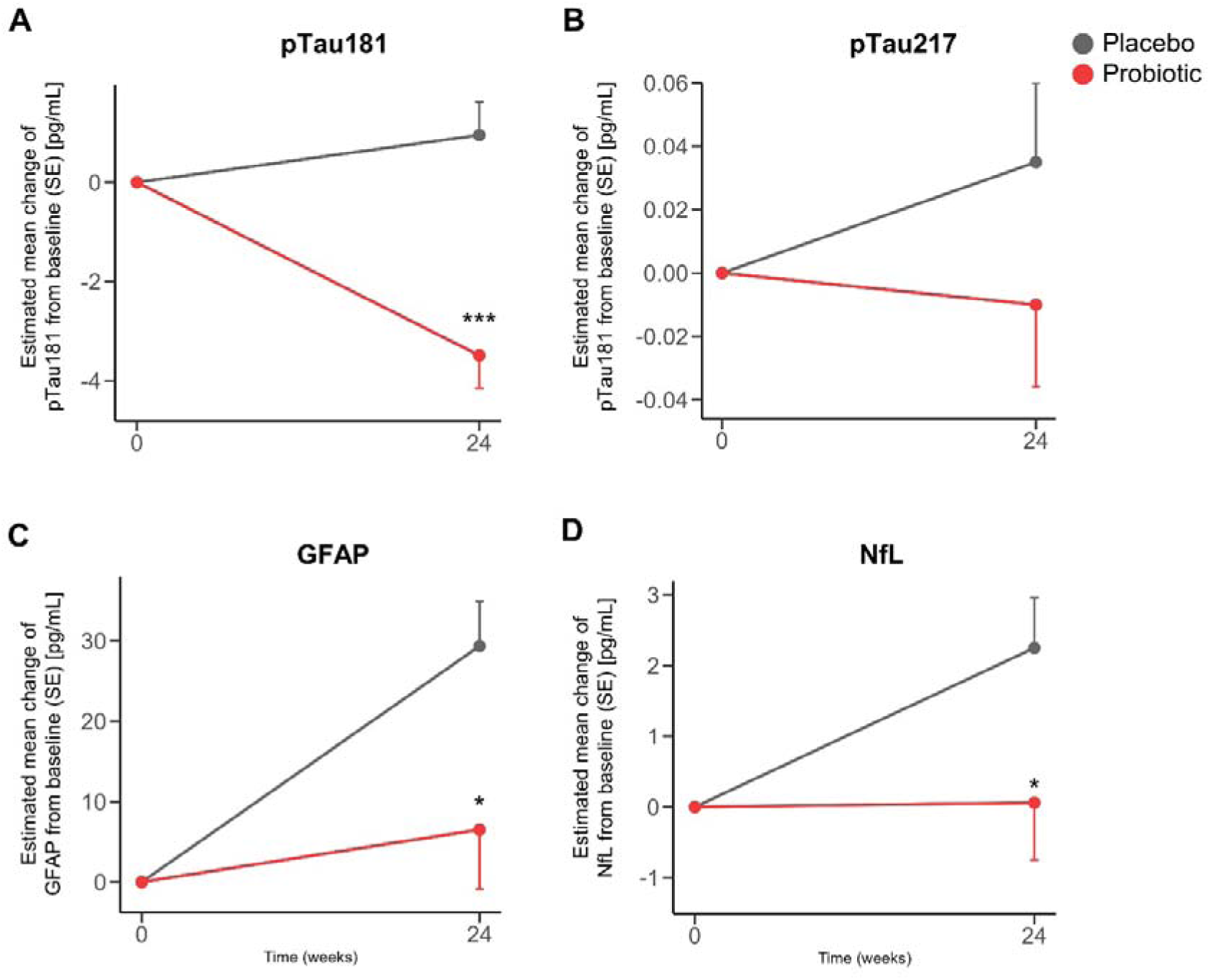
Changes in plasma biomarkers in the probiotic and placebo groups. Line graph (A-D) showing the change from baseline to 24-weeks in pTau181, pTau217, GFAP, and NfL. The lines represent mean change values relative to baseline, with placebo shown in black and probiotic shown in red; error bars indicate standard errors. Group differences are annotated using significance (*p<0.05, **p<0.01, ns=not significant). Abbreviations: pTau: phosphorylated tau; GFAP: glial fibrillary acidic protein; NfL: neurofilament light chain;

**Table 3.**
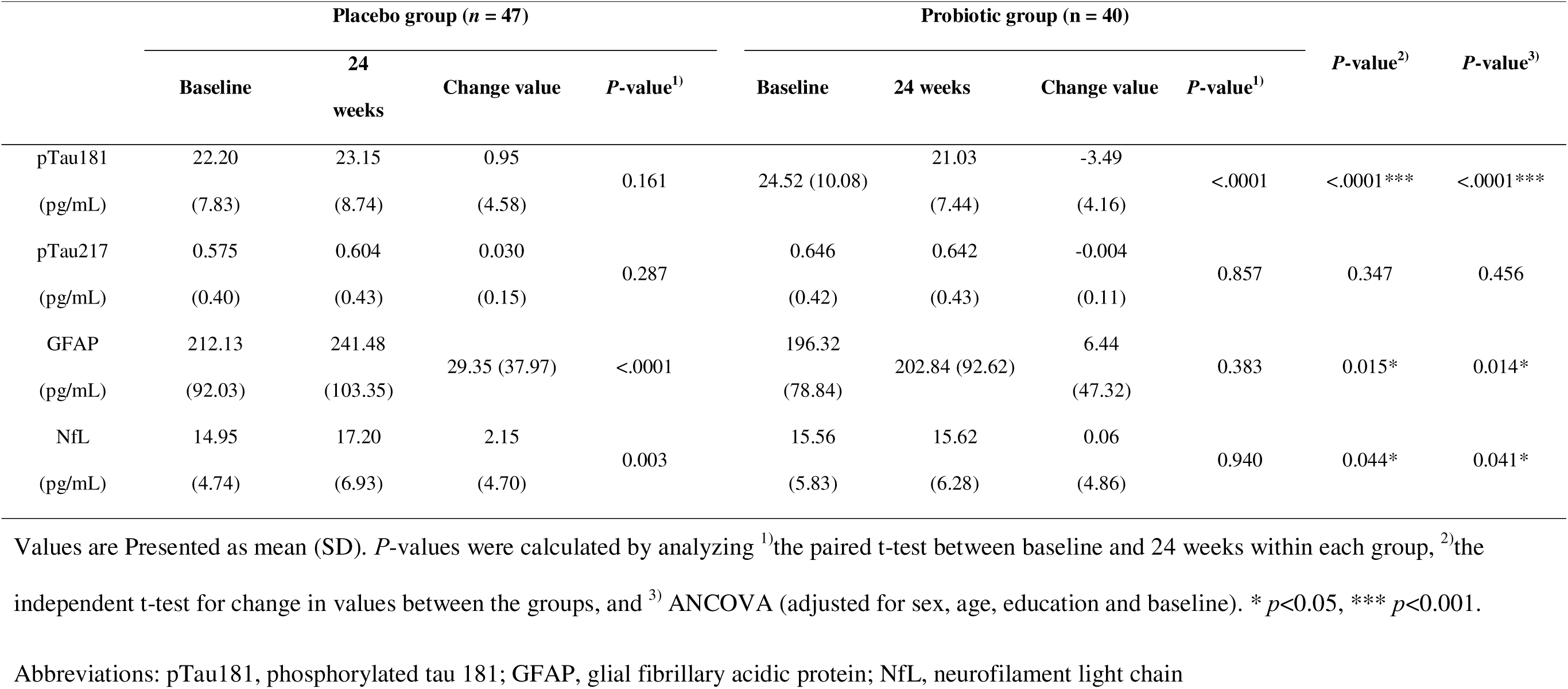
Group difference in biomarkers of Alzheimer’s disease.

## Discussion

We investigated the clinical efficacy of a multistrain probiotic supplement targeting the oral microbiota on Alzheimer’s disease (AD)-related biomarkers and cognitive function in older adults with early cognitive impairment. In this 24-week, double-blind, randomized, placebo-controlled trial, the administration of the multistrain probiotic resulted in significant improvements in both plasma-based AD biomarkers and domain-specific cognitive performance. Most notably, probiotic supplementation was associated with a marked reduction in plasma pTau181 levels. This effect was not observed in the placebo group and occurred alongside the preservation of visuospatial construction and visual memory abilities. Given the increasing consensus that targeting modifiable biological systems is most effective during the prodromal stage of AD, our findings provide converging biomarker and neuropsychological evidence to support the modulatory role of microbial interventions in early neurodegenerative processes.

Overall, we found that a multistrain probiotic supplement was highly effective at positively modulating key AD biomarkers. Most notably, compared with the placebo group, the probiotic group showed a significant reduction in plasma pTau181, a critical marker of tau pathology in the early stages of AD [22, 23]. Phosphorylated tau species are strongly associated with tau aggregation, disease severity, and cognitive decline [1]. Therefore, the observed reduction is noteworthy in the context of the 24-week intervention. Furthermore, the stabilization of GFAP and NfL levels, compared with their significant increase in the placebo group, is indicative of attenuated neuroinflammatory and neurodegenerative signaling. These biomarker patterns aligned with the established actions of the probiotic species included in the formulation. Overall, *Bifidobacterium animalis ssp. lactis* supports a healthier oral environment by strengthening the epithelial barrier and lowering systemic inflammatory signaling, thereby reducing inflammatory inputs to the brain [24]. Prior studies have further demonstrated that *Limosilactobacillus fermentum* exerts strong anti-inflammatory effects, including suppression of IL-1β, TNF-α, and nitric oxide production, thereby attenuating inflammation [14, 25]. *Levilactobacillus brevis* strains also demonstrated biologically relevant properties; they are recognized for their GABA-producing and antioxidant properties, which may reduce oxidative stress, a known promoter of pathological tau phosphorylation [26]. These strain-specific biological activities provide a coherent mechanistic explanation for the reduction in pTau181 and stabilization of injury-related biomarkers in the probiotic group. These results are consistent with prior animal studies showing that probiotic administration can diminish amyloid-β deposition, suppress tau phosphorylation, and modulate microglial activity [6, 8].

Clinically, the probiotic group demonstrated a significant improvement in the global CDR; this effect was specifically traced to the CDR memory domain, which reflects everyday memory function and distinguishes it from the neuropsychological test performance. Given that the placebo group showed no comparable changes, this finding suggests a possible functional advantage of probiotic intake during the intervention period.

Regarding neuropsychological function, the probiotic group demonstrated significant improvements in visuospatial construction tasks (CERAD Constructional Praxis and ROCF copy), whereas the placebo group showed no such improvement. Moreover, constructional recall remained stable in the probiotic group but declined significantly in the placebo group, with a robust group × time interaction. Visuospatial recall, as measured by the CERAD CR, is considered dependent on context-free memory, which involves the retrieval of information without reliance on contextual cues and is less scaffolded by meaning or verbal strategies. Context-free memory has been reported to be sensitive to very early vulnerabilities in the posterior cortical network along the AD continuum [27]. In contrast, verbal memory and context-dependent memory improved in both groups without a between-group difference. This pattern is most parsimoniously explained by the practice effects that commonly accompany serial neuropsychological testing, particularly on semantically supported verbal tasks. This divergence is consistent with prior evidence showing that visuospatial and visual episodic memory functions are particularly sensitive to tau pathology [10–13], supporting the biological plausibility of a tau-linked cognitive benefit in this domain. Visuospatial memory declines early and sensitively track tau accumulation, distinguishing it from verbal learning tasks, which are more susceptible to retest effects. Notably, the pattern of cognitive preservation in the probiotic group mirrored the biomarker findings: reductions in pTau181 and stabilization of GFAP/NfL occurred alongside improved or preserved visuospatial construction and visual memory, whereas the placebo group exhibited both biomarker worsening and poorer trajectories in visuospatial tasks. This convergence is biologically plausible given the mounting evidence that oral-immune–brain axis mechanisms, including immune modulation, microglial regulation, and altered metabolite signaling, can shape the neuroinflammatory and neurodegenerative pathways relevant to AD [3, 4]. The specific patterns of change, most notably the reduction in pTau181, together with the selective improvements in visuospatial cognition, suggest that probiotic intervention may interact with the neurobiological pathways that are vulnerable in the very early stages of AD. Further studies are, therefore, required to elucidate the underlying mechanisms. The precise mechanisms linking oral-derived immune or metabolic signaling to tau-related processes and their associated cognitive consequences remain unclear, underscoring the need for additional research to clarify these complex interactions.

Meanwhile, executive and attention functions, measured by the TMT and Stroop test, remained unchanged in both groups. Because our participants were all in the very early stage of cognitive impairment, executive functions, particularly those mediated by the frontal systems, may tend to be relatively preserved in the very early stages of Alzheimer’s disease [28]. This cognitive domain may require longer exposure times or more sensitive measures to detect subtle changes.

## Limitations

Despite its important implications, this study has several limitations that warrant consideration and provide directions for future research. First, the sample size was relatively small, which may have limited the statistical power to detect subtle treatment effects, particularly in cognitive domains that show modest changes over short intervals. Therefore, a larger sample size is required for replication. Second, the 24-week intervention period, although sufficient to detect changes in plasma biomarkers, may have been too short to capture long-term functional changes in cognition. Longer trials are required to determine the durability and clinical relevance of these effects, and we are currently conducting follow-up studies. Third, although neuropsychological testing was conducted following standardized procedures, repeated assessments may have introduced practice effects, particularly in verbal tasks that rely on semantic cueing. Future studies may benefit from alternative test forms or cognitive paradigms less susceptible to retest effects. Finally, the study did not include follow-up imaging of tau or Aß PET, limiting the ability to directly relate changes in plasma biomarkers to in vivo brain pathology. Integrating plasma biomarkers with PET imaging and MRI in future trials will allow a more definitive assessment of whether probiotic-related changes reflect the true modulation of AD pathology; indeed, we are currently incorporating neuroimaging in ongoing long-term follow-up studies.

## Conclusions

In conclusion, our findings from this 24-week, double-blind, randomized, placebo-controlled clinical trial suggest that multistrain probiotic supplementation targeting the oral microbiota may beneficially modulate tau-related biological pathways and exert selective cognitive benefits in domains strongly associated with early tau pathology. The parallel reduction in plasma pTau181 and the preservation of visuospatial construction and visual memory provide converging evidence supporting the potential role of microbiota–immune–brain interactions in shaping early neurodegenerative trajectories. While these findings are preliminary, they should motivate larger, longer trials incorporating tau/Aβ PET imaging to verify the durability of these effects and to test whether visuospatial memory serves as a sensitive clinical endpoint for neuroprotective interventions. Critically, the observed positive effects on both biomarkers and cognition underscore the significant potential of this specific multistrain probiotic supplement (*Bifidobacterium animalis ssp lactis KL101, Limosilactobacillus fermentum KL271, and Levilactobacillus brevis KL251*) as a readily available, non-pharmacological strategy for the management of the earliest stages of Alzheimer’s disease.

## Data Availability

The datasets analyzed in the current study are not publicly available but can be obtained from the corresponding author upon reasonable request.

## Abbreviations

AD: Alzheimer’s disease;
APOE: apolipoprotein E;
Aß: Amyloid-beta;
CDR: a clinical dementia rating;
CN: cognitively normal;
CSF: cerebrospinal fluid;
GARD: the Gwanju Alzheimer’s Disease and Related Dementia;
MMSE: Mini-Mental State Examination;
CERAD: Consortium to Establish a Registry for Alzheimer’s Disease;
MCI: mild cognitive impairment;
MRI: magnetic resonance imaging;
NFT: neurofibrillary tangles;
PET: positron emission tomography;
p-Tau: phosphorylated tau;
GFAP: glial fibrillary acidic protein;
NfL: neurofilament light chain;
ROCF copy: the Rey-osterrieth complex figure test copy;
ROCF delayed: the ROCF immediate recall 20-minute delayed recall;
ROCF imm: the ROCF immediate recall;
GDS: Geriatric Depression Scale;
SCWT: Stroop Color and Word Test
VF: Verbal fluency;
BNT: Boston Naming Test;
WLM: Word List Memory;
WLR: Word List Recall;
WLRc: Word List Recognition;
CP: Constructional Praxis
CR: Constructional Recall;
TMT: trail making tests

## Declarations

### Ethics approval and consent to participate

This study was approved by the Institutional Review Board of Chonnam National University Hospital (IRB no: CNUH-2023-072). Written informed consent was obtained from each participant or their legal guardian. The study was conducted in accordance with the principles of the Declaration of Helsinki.

### Consent for publication

Not applicable

### Authors’ contributions

KHL and KHC designed the research. SGK and KHC collected and reviewed the data. EHS and SRK undertook the statistical analysis. KHL, EHS, SRK, HJY, KYC, SGK, and KHC interpreted the data. EHS and SRK wrote the draft of the manuscript. KYC, HJY, SGK, KHL, and KHC critically revised the manuscript. All authors have approved the submitted version.

## Acknowledgements

Not applicable

